# What Role Should EBU Catheters Play in the Interventional Approach to Anomalous Right Coronary Arteries?

**DOI:** 10.1101/2025.06.03.25328934

**Authors:** Mikias Legesse Gebremedhin, Milan Sigdel, Zhao Ruixue, BinBin Du, Yanzhou Zhang

**Affiliations:** Department of Cardiology, The First Affiliated Hospital of Zhengzhou University, Zhengzhou, China; Department of Intervention Radiology, The First Affiliated Hospital of Zhengzhou University, Zhengzhou, China

**Author notes:** Corresponding Author - Yanzhou Zhang, Department of Cardiology, The First Affiliated Hospital of Zhengzhou University, Mianfang St, Erqi District, Zhengzhou, Henan, 450000, China. Co-Author Contact Details. **Ethics & Integrity Statements**. **Ethics Approval** This retrospective study was approved by the Institutional Ethical Review Board of The First Affiliated Hospital of Zhengzhou University (IRB No. 2024-KY-0118-272). **Patient Consent** Requirement for individual patient consent was waived by the Institutional Ethical Review Board dee to the retrospective nature of the study. **Funding Statement** This work received no external funding. **Permission to Reproduce Material** No previously published figures, tables, or illustrations were reproduced in this manuscript. **Clinical Trial Registration** Not applicable.

**Keywords:** Anomalous coronary Artery, Percutaneous Coronary Interventions, EBU Coronary catheters, Anomalous RCA

## Abstract

**Background:** Percutaneous coronary intervention (PCI) for anomalous right coronary artery (ARCA) remains technically challenging due to variable ostial anatomy. The Extra Back-Up (EBU) catheter, although designed for left coronary interventions, may offer advantages in ARCA PCI, but its performance across anatomical subtypes is not well defined.

**Methods:** This single-center retrospective study included 17 patients (2019–2024) with ARCA from the left coronary sinus or with high take-off anatomy, in whom an EBU catheter was used. Fourteen patients underwent PCI, one had FFR-only assessment, and two underwent diagnostic angiography alone. Anatomical subtypes were: Type A (ostium above left coronary artery [LCA]), Type B (below LCA), Type C (midline), or high take-off (≥10 mm above sinotubular junction). Primary outcomes were procedural success (stable engagement, full device delivery, no catheter exchange) and safety.

**Results:** Procedural success with the EBU catheter was 100% in Type A (2/2) and 83.3% in Type B (5/6) when used as the initial guide. In Type C and high take-off anatomies, success was achieved in one of two cases each. EBU was also employed as a second-choice catheter in five patients after failed engagement with standard guides catheters. No catheter-related dissections occurred. One minor in-hospital complication (5.9%) and one patient had unplanned Re-intervention within 30 days.

**Conclusions:** EBU catheters are effective in ARCA PCI, especially in Type A and B anatomies, but less so in Type C and high take-off variants. These findings support anatomy-tailored guide selection. Multicenter studies are needed to further define role of EBU in this subset.

## Introduction

Anomalous coronary arteries represent a spectrum of congenital variations that pose unique challenges for percutaneous coronary intervention (PCI)[1][2][3]. Anomalous coronary artery from the opposite sinus (ACAOS) is a rare congenital condition, affecting approximately 0.2–2.0% of the population[4][5]. Among these, anomalous origin of the right coronary artery (ARCA) from the left sinus of Valsalva (LSOV) constitutes a particularly complex subgroup, with an estimated prevalence of 0.26-0.5% in angiographic studies[6][7]. The clinical significance of these anomalies stems from their association with myocardial ischemia, arrhythmias, and sudden cardiac death, particularly in young athletes during physical exertion[8][9].

Anatomically, ARCA from LSOV can be classified into three main subtypes 6(Figure 1) based on ostial location relative to the left coronary artery (LCA)mirrored from previous study[10]:

**Figure 1:**
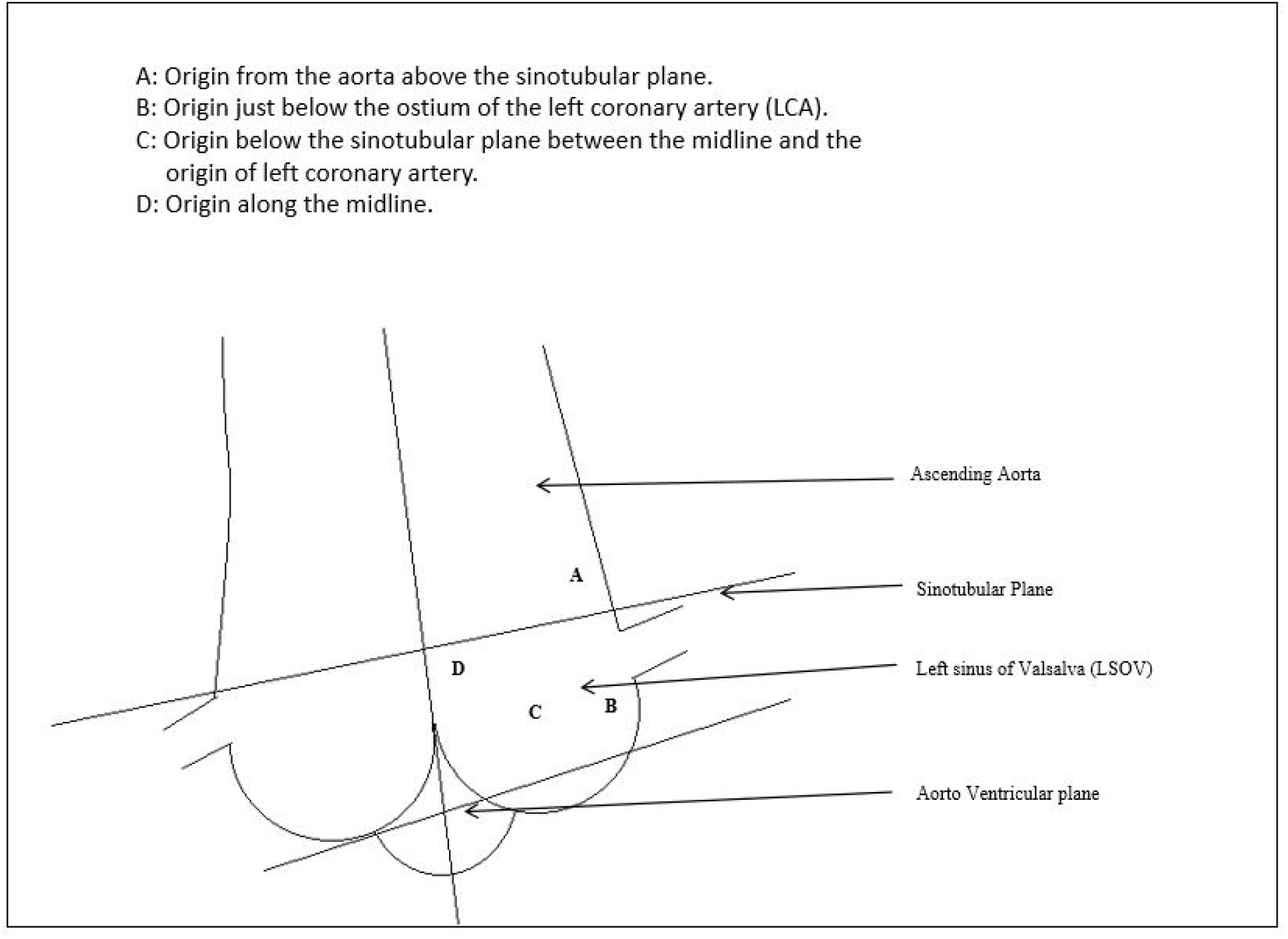
A simple anatomical illustration showing anomalous RCA classification according to ostium location (relation to sino-tubular plane and Left main ostium)

Type A: Ostium positioned above the LCA (6-27% of cases)

Type B: Ostium located below the LCA (most common variant)

Type C: Midline origin between the LCA and aortic midline

Additionally, high take-off anomalies (>1 cm above the sinotubular junction) represent a distinct entity with prevalence estimates of 0.006-0.2%[11]

Current PCI strategies for ARCA-LSOV rely heavily on operator experience and ad hoc catheter selection. While conventional guides (Judkins Right, Amplatz Left) are commonly used in standard cases[12], their efficacy decreases for anomalous origin from LSOV and high take-off anomalies[7]. This has led to exploration of alternative approaches, including radial-dedicated catheters (Ikari Left) and left coronary guides repurposed for right coronary use [13].

The Extra Back-Up (EBU) catheter, originally designed for left coronary interventions, offers several theoretical advantages for ARCA-LSOV PCI, including enhanced passive support from contralateral aortic wall contact [14],], a wire-modifiable curvature that allows adaptation to various takeoff angles, and a single-catheter strategy that may reduce the need for equipment exchanges

Despite these potential benefits, systematic evaluation of EBU catheters in ARCA-LSOV PCI remains lacking. This study analyzes our center’s experience with EBU-guided PCI in 17 consecutive cases of ARCA, focusing on procedure success rates across anatomical subtypes, frequency of catheter exchanges, and safety outcomes including complication rates. Our findings aim to establish an evidence-based algorithm for catheter selection in these challenging anatomies, potentially simplifying the interventional approach while maintaining safety and efficacy.

## Methods

### Study Design and Setting

This retrospective study included all patients who underwent coronary angiography, with or without PCI, between May 2019 and June 2024 at a tertiary care center. From this group, we selected patients with an anomalous right coronary artery (ARCA) in whom an EBU catheter was used for either angiography or PCI.

The study was approved by the Institutional Ethics Committee of the First Affiliated Hospital of Zhengzhou University (IRB No. 2024-KY-0118-272). Due to its retrospective design, the requirement for informed consent was waived. Medical records were accessed for research purposes between July and August 2024. All data were fully anonymized before access, and no identifiable patient information was available to the authors at any stage of data collection or analysis.EBU was chosen either as the initial guide catheter (n=12) or after unsuccessful attempts with other catheters (n=5).

Seventeen cases met inclusion criteria: 14 underwent PCI, one had FFR assessment, and two were diagnostic-only procedures. Cases in which EBU was not attempted were excluded.

### Anatomical Classification

ARCA-LSOV cases were categorized into three anatomical subtypes based on angiographic origin relative to the LCA ostium[10]:

Type A: Ostium above the LCA

Type B: Ostium below the LCA

Type C: Midline or commissural origin between the LCA and right sinus

An additional group included patients with high take-off RCA, defined as origin ≥10 mm above the sinotubular junction.

### Procedure Protocol

All procedures were performed via transradial access. Operators selected EBU (typically 3.5 or 4.0) based on ostial direction and procedural goals. Adjunctive techniques such as guide extension or microcatheters were employed at operator discretion.

### Data Collection and Endpoints

Patient demographic data, anatomical classification, and procedural details were collected from institutional records and catheterization reports.

Key outcomes included:

Primary Endpoint: Procedural success with EBU catheter, defined as successful RCA engagement, full device delivery, and procedure completion without catheter exchange.

Secondary Endpoints

- Need for guide catheter exchange
- Use of guide extension or adjunctive tools
- Catheter-related complications (e.g., ostial dissection, pressure damping)
- Clinical adverse events within 30 days

### Ethical Approval

The study was approved by the Institutional Ethical Review Board of the First Affiliated Hospital of Zhengzhou University (IRB No. 2024-KY-0118-272), and all data were handled in accordance with relevant ethical guidelines and patient confidentiality standards.

## Results

### Patient and Anatomic Characteristics

Seventeen patients with angiographically confirmed anomalous right coronary artery (ARCA) underwent coronary angiography or PCI using an EBU catheter. The mean age was 61.3 ± 9.8 years, and 70.6% were male. Hypertension was the most common comorbidity (58.8%), followed by a history of smoking (41.2%), diabetes mellitus (17.6%), and chronic kidney disease (11.8%). Most patients presented with stable angina (58.8%), while 23.5% had non–ST elevation acute coronary syndrome; no cases of STEMI were recorded. Three patients (17.6%) underwent diagnostic angiography or FFR-only assessment without intervention (Table 1).

**Table 1.** Baseline Clinical Characteristics of Patients Undergoing ARCA with EBU Catheter.

| Characteristic | n (%) |
| --- | --- |
| Total patients | 17 (100%) |
| Male sex | 12 (70.6%) |
| Female sex | 5 (2.9%) |
| Age, mean ± SD (years) | 61.3 ± 9.8 |
| Hypertension | 10 (58.8%) |
| Diabetes mellitus | 3 (17.6%) |
| Chronic kidney disease | 2 (11.8%) |
| Current or former smoker | 7 (41.2%) |
| Clinical presentation |  |
| – Stable angina | 10 (58.8%) |
| – Non-ST elevation ACS | 4 (23.5%) |
| – STEMI | 0 (0%) |
| – Diagnostic or FFR only | 3 (17.6%) |

The majority of ARCA cases (82.4%) originated from the left coronary sinus; classified as Type A, B, or C; while the remaining 17.6% had high take-off anatomy, defined as an origin ≥10 mm above the sinotubular junction (Table 2).

**Table 2.** Anatomic and Procedural Features of ARCA Cases Treated with EBU Catheter.

| Characteristic | n (%) |
| --- | --- |
| RCA origin from left cusp (Type A–C) | 14 (82.4%) |
| – Type A (above LCA ostium) | 3 (17.6%) |
| – Type B (below LCA ostium) | 9 (52.9%) |
| – Type C (midline/commissural origin) | 2 (11.8%) |
| High take-off RCA ($\geq 10$ mm above STJ) | 3 (17.6%) |
| Chronic total occlusions (CTO) | 2 (11.8%) |
| Radial access used | 17 (100%) |
| EBU used as first-choice catheter | 12 (70.6%) |
| Use of guide extension catheter | 7 (41.2%) |

### EBU Catheter Use and Procedural Outcomes

The EBU catheter was used as the first-choice guide in 12 patients, with varying success depending on RCA origin type (Table 3). Success was defined as stable engagement, device delivery, and completion of the procedure without catheter exchange.

**Table 3.** EBU Catheter Performance and Usage Sequence by ARCA Subtype.

| **Subtype** | **Total Cases** | **EBU**<br>**Used as**<br>**1st**<br>**Choice** | **Successful**<br>**with EBU** | **Converted**<br>**from EBU** | **Extension**<br>**Used** | **EBU Used After**<br>**Failed Initial Catheter** |
| --- | --- | --- | --- | --- | --- | --- |
| Type A | 3 | 2 | 2 | 0 | 1 | 1 (initial JR,JL) |
| Type B | 9 | 6 | 5 | 1 (to JL) | 3 | 3 (initial, AL) |
| Type C | 2 | 2 | 1 | 1 (to AL) | 2 | 0 |
| High<br>Take-Off | 3 | 2 | 1 | 1 (to SAL) | 1 | 1 (initial:JR,JL) |
| **Total** | **17** | **12** | **9** | **3** | **7** | **5** |

- **Type A (n=3):** EBU used first in 2 cases and successfully in both. One additional case used EBU after a failed initial catheter. One patient required guide extension support.
- **Type B (n=9):** EBU used as first catheter in 6 cases, with 5 successful. One case required conversion to Judkins Left. In three additional B-type cases, EBU was used after failure of other catheters. Three patients required guide extension support. Representative pre- and post-intervention angiographic images of a Type B anomalous RCA PCI using EBU as shown in Figure 2.
- **Type C (n=2):** EBU used first in both cases; one succeeded one required conversion to Amplatz Left. Both cases used guide extension catheters.
- High Take-Off (n=3): EBU used first in 2 cases, with one success and one requiring conversion. One additional case used EBU after a failed attempt with another catheter. One case used guide extension support.

**Figure 2:**
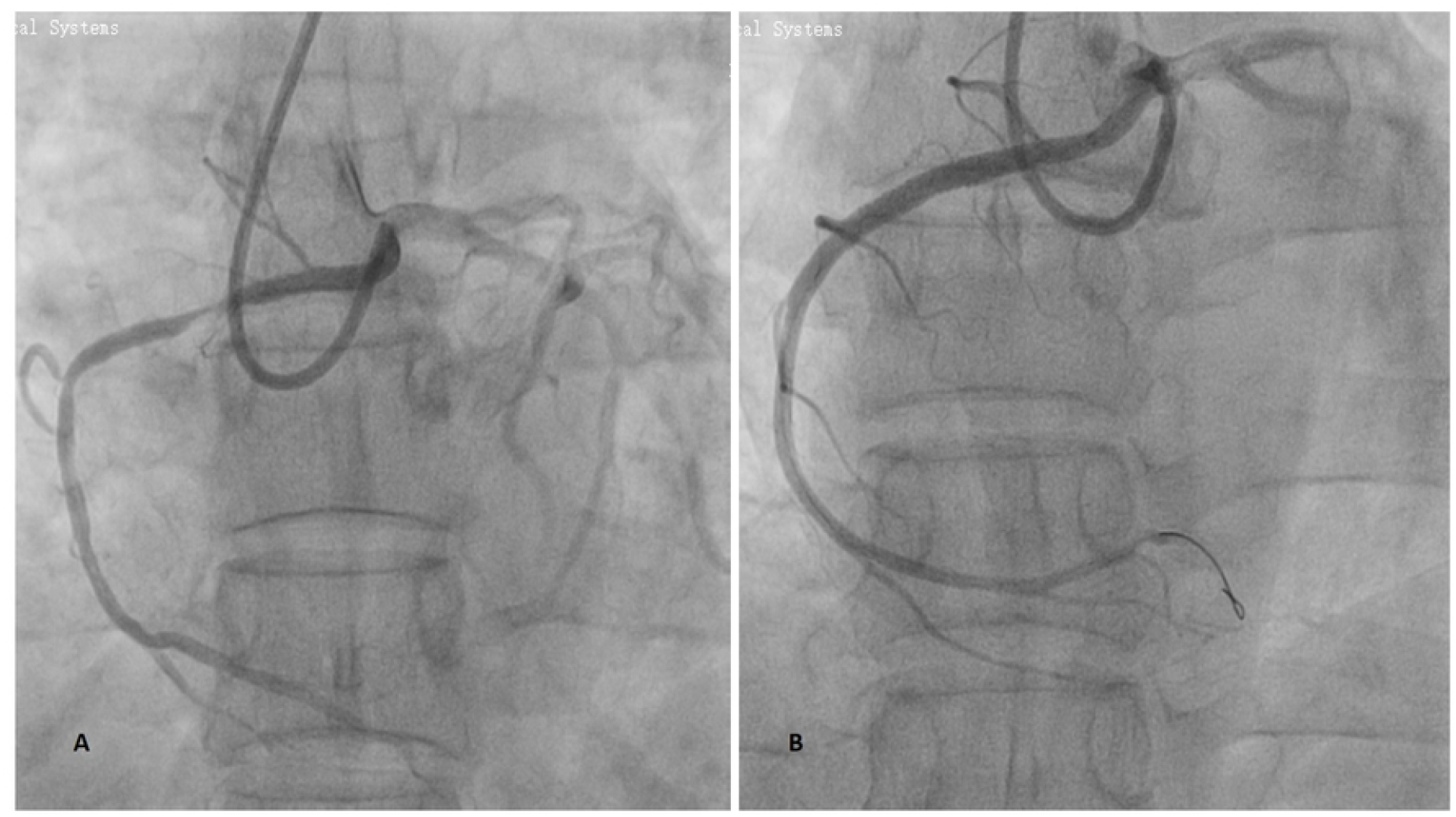
**A-** Angiographic image of Type B anomalous RCA by EBU guide catheter with ostium originating just below the left main ostium showing a proximal to mid segment stenotic lesion**; B-** Post PCI procedure image showing satisfactory outcome after utilizing EBU guide catheter for the procedure

### Procedural Safety

No catheter-induced coronary dissections were observed. One patient experienced a minor in-hospital complication unrelated to catheter selection. One patient required re-intervention PCI for revascularization within thirty postoperative days (Table 4).

**Table 4.** Intra and Post operative Complications.

| **Variable** | **n (%)** |
| --- | --- |
| Catheter-induced dissection | 0 (0%) |
| In-hospital complications | 1 (5.9%) |
| Unplanned Re-intervention within 30 days | 1 (5.9%) |

## Discussion

Percutaneous intervention in ARCA presents distinct technical challenges due to ostial eccentricity, unusual take-off angles, and variable alignment with standard catheter geometries. This case series explored the procedural utility of the EBU catheter; originally designed for left coronary interventions, in a consecutive series of ARCA-LSOV cases.

Our findings suggest that the EBU catheter may serve as a viable first-choice option in select anatomical subtypes. Specifically, procedural success was highest in Type A (100%) and Type B (83.3%) anatomies when EBU was used as the initial guide. In these cases, the natural leftward orientation may facilitate better alignment with the EBU’s curve. However, EBU performance was less favorable in Type C (50%) and high take-off (50%) anatomies, where guide stability or coaxiality was more difficult to achieve..

These observations align with prior studies describing the technical complexity of ARCA-LSOV PCI. For instance, Sarkar et al. emphasized shape-specific catheter matching based on ostial origin using FL, FCL, VL, and AL curves tailored to take-off type[10]. Nevertheless, a stepwise and flexible approach to catheter selection remains essential, allowing operators to escalate as needed based on procedural challenges.

The design of the EBU catheter may offer a mechanical advantage in ARCA cases where the origin lies near the left coronary cusp, particularly in Type A and B configurations. Unlike standard right coronary catheters, the EBU’s curve is optimized for left coronary engagement, providing a natural alignment with ARCAs that originate close to the LCA ostium. This geometry allows for better coaxial engagement in ostia with an anterior or superior trajectory relative to the aorta. Additionally, the EBU’s strong passive support from the contralateral aortic wall can help stabilize the guide, which is critical in tortuous or high-resistance lesions. These structural characteristics explain the high procedural success observed in our Type A and B cases, where the catheter’s alignment and backup support synergized with the RCA take-off angle, reducing the need for multiple exchanges or aggressive manipulation.

This study aims to identify the scenarios in which the EBU catheter is most effective for ARCA interventions, without promoting a uniform EBU-first strategy. Since performance of other catheters in these cases was not analyzed, the findings are intended to add to the procedural literature and provide real-world insights for interventionists using the EBU catheter in ARCA cases.

In conclusion, while the EBU catheter shows promising effectiveness in specific anatomical subtypes of ARCA, particularly those with origins near the left coronary ostium, its utility is limited in others. This underscores the need for tailored catheter selection based on anatomical characteristics rather than a one-size-fits-all approach. By detailing the strengths and limitations of the EBU catheter in real-world practice, this study contributes valuable insights to guide interventionists managing the technical challenges of ARCA PCI.

## Strength and limitations

Our cohort represents a selective but clinically relevant population; ARCA cases where operators judged the EBU catheter to offer technical advantages. While this excludes cases successfully managed with other catheter types, the five-year collection period and case volume are consistent with the rarity of this anomaly in tertiary centers. Nonetheless, the study is limited by its single-center, retrospective design, selective inclusion of EBU cases, and modest sample size (n=17), which restrict generalizability and limit subgroup analysis. Anatomical classification relied on operator assessment without core lab validation or advanced imaging, and follow-up was confined to procedural and short-term outcomes. Despite these limitations, the study provides meaningful procedural insights into the use of EBU catheters across varied anatomical subtypes.

## Data Availability

Data will be available from the corresponding author upon reasonable request.

## Notes

**Data Availability** The data that support the findings of this study are available from the corresponding author upon reasonable request.

**Conflict of Interest Disclosure** The authors declare no conflicts of interest related to this study.

### Competing Interest Statement

The authors have declared no competing interest.

### Funding Statement

The author(s) received no specific funding for this work.

### Author Declarations

This retrospective study was approved by the Institutional Ethical Review Board of The First Affiliated Hospital of Zhengzhou University (IRB No. 2024-KY-0118-272).

